# Impact of Convalescent Plasma Transfusion (CCP) In Patients With Previous Circulating Neutralizing Antibodies (nAb) to COVID-19

**DOI:** 10.1101/2020.12.08.20246173

**Authors:** Ana Paula H Yokoyama, Silvano Wendel, Carolina Bonet-Bub, Roberta M Fachini, Ana Paula F Dametto, Fernando Blumm, Valeria F Dutra, Gabriela Candelaria, Araci M Sakashita, Rafael Rahal Guaragna Machado, Rita Fontao-Wendel, Nelson Hamerschlak, Ruth Achkar, Murillo Assuncao, Patricia Scuracchio, Victor Nudelman, Laerte Pastore, Joao R R Pinho, Mirian Dal Ben, Roberto Kalil Filho, Alexandre R. Marra, Mariane T. Amano, Esper Kallas, Alfredo Salim Helito, Carlos Roberto Ribeiro de Carvalho, Danielle Bastos Araujo, Edison Luiz Durigon, Anamaria A Camargo, Luiz V Rizzo, Luiz F Lima Reis, Jose M Kutner

**Affiliations:** Hospital Israelita Albert Einstein, Sao Paulo, Brazil; Hospital Sirio-Libanes Blood Bank, Sao Paulo, Brazil; Hospital Sirio-Libanes, Sao Paulo, Brazil; Hospital Sirio-Libanes, Brasilia, Brazil; Office of Clinical Quality, Safety, and Performance Improvement, University of Iowa Hospitals and Clinics, Iowa City, Iowa; Department of Microbiology, Institute of Biomedical Sciences, University of Sao Paulo, Sao Paulo, Brazil; University of Sao Paulo – Heart Institute (Incor), Sao Paulo, Brazil; Department of Infectious and Parasitic Diseases, School of Medicine, University of Sao Paulo, Sao Paulo, Brazil; Cardio-Pulmonary Department, Pulmonary Division, Heart Institute (Incor), University of Sao Paulo, Sao Paulo, Brazil; Scientific Platform Pasteur USP, Sao Paulo, Brazil

## Abstract

**INTRODUCTION:** COVID-19 convalescent plasma (CCP) transfusion has emerged in the past months as an alternative approach to treat pneumonia cases of SARS-CoV-2. Current evidence regarding characteristics of the plasma product, the titer of neutralizing antibodies (nAbs) in the transfused units, time to onset of intervention, and impact of nAbs produced by the patient are limited and heterogeneous.

**MATERIAL AND METHODS:** We describe the preliminary results of 104 patients with severe pneumonia due to SARS-CoV-2 transfused with CCP at three medical centers in Brazil. All enrolled patients were transfused with doses between 200 mL through 600mL of ABO compatible CCP on days 0-2 after enrolment. Clinical parameters were monitored and nAbs titration was performed using the cytopathic effect-based virus neutralization test with SARS-CoV-2 (GenBank MT126808.1).

**RESULTS:** Forty-one patients achieved clinical improvement on day 14, and multivariable logistic regression showed that nAbs T (from CCP units transfused) (p= 0.001), nAbs P0 (on day of enrolment) (p=0.009) and use of other supportive therapies (p<0.001) were associated with higher odds for this clinical improvement. Considering ICU length of stay (LOS) and length of mechanical ventilation, in our analysis, nAbs P0 were associated with a significant reduction in ICU LOS (p=0.018) and duration of mechanical ventilation (p<0.001). Administration of CCP after 10 days of symptom onset was associated with increases in ICU length of stay (p<0.001).

**DISCUSSION/CONCLUSION:** Despite the study limitations, our data have shown an association between patients’ previously acquired nAbs and clinical outcomes. The potential value of timely administration of CCP transfusion before day 10 of disease onset was demonstrated and nAbsP0, but not nAbsT, were associated with ICU LOS, and duration of mechanical ventilation on the improvement of clinical outcomes was also demonstrated. In conclusion, we consider these data are useful parameters to guide future CPP transfusion strategies to COVID-19.

## Introduction

COVID-19 convalescent plasma (CCP) transfusion has emerged in the past months as an alternative approach to treat pneumonia cases of Severe Acute Respiratory Syndrome Coronavirus 2 (SARS-CoV-2) [1,2]. Several specific therapies are still under development, with limited evidence towards efficacy and survival benefit [3,4,5]. Hence, given its rapid availability, CCP transfusion was proposed as an alternative therapy early in the course of Coronavirus 2019 (COVID-19) pandemics, with proven safety [6]. Historically, convalescent plasma has been used as an emergent strategy in many outbreaks, such as Spanish influenza, with reduced mortality rates [7]. Positive results have been described in other viral epidemics like Ebola [8], Severe Acute Respiratory Syndrome for Coronavirus (SARS-CoV-1) [9], and Middle East Respiratory Syndrome (MERS-CoV) [10].

The rationale for CCP transfusion relies on the fact that convalescent individuals have circulating neutralizing antibodies (nAbs) able to suppress the infection [11]. Other immunomodulatory mechanisms, such as complement activation, antibody-dependent cytotoxicity, or phagocytosis are potential pathways through which CCP might alleviate systemic inflammation. Also, non-nAbs that bind to the virus, despite not interfering with replication capacity, might contribute to recovery [12]. Previous studies demonstrated that CCP transfusion was able to inhibit SARS-CoV-2 replication [13] and efficacy was associated with the concentration of nAbs in the plasma of convalescent donors [14]. However, data about the clinical impact of nAbs on COVID-19 outcomes are still sparse.

Current evidence regarding characteristics of the plasma product, the titer of nAbs in the transfused units, time to onset of intervention, and impact of nAbs produced by the patient are limited and heterogeneous, which precludes consistent conclusions from the available data. We describe here the preliminary results of 104 patients with severe pneumonia due to SARS-CoV-2 transfused with CCP at three medical centers in Brazil.

## Materials and methods

This was a single-arm prospective study conducted at three reference medical centers in Sao Paulo and Brasilia, Brazil. It was approved by the Brazilian National Commission for Research Ethics (CONEP), CAAE 30922420.6.2002.0071, and the Ethics Committees of each participating site. Informed Consent was obtained from all study participants or their legal representatives.

### Convalescent plasma collection

CCP donors with laboratory-confirmed COVID-19 diagnosis by reverse transcriptase-polymerase chain reaction (RT-PCR), fully recovered and asymptomatic for at least 14 days were recruited on participating sites. Donor screening comprised Brazilian regular criteria for blood donation and negative RT-PCR from blood or naso-oropharyngeal swabs collected on the day of donation. Serum for determination of SARS-CoV-2 nAbs from each donor was also collected on the day of donation for further analysis. Convalescent plasma products were obtained from whole blood or plasmapheresis donations [15], and approximately 50% of units were pathogen-inactivated using amotosalen/UVA illumination (INTERCEPT, Cerus corporation, Concord, CA USA).

### Patients

From April 11 through October 25, 104 patients admitted to participating sites with COVID-19 confirmed diagnosis by RT-PCR, &[gt]18 years of age, with criteria for severe pneumonia (defined by respiratory distress: oxygen saturation of 93% or less on room air, respiratory rate > 30 breaths/min and/or arterial partial pressure of oxygen (PaO_2_) /fraction of inspired oxygen (FiO_2_) of 300 or less) were included. Patients with preexisting history of anaphylactic transfusion reaction, pregnant or lactating women were excluded.

### Intervention

All enrolled patients were transfused with doses between 200 mL through 600mL of ABO compatible CCP on days 0-2 after enrolment, with close monitoring. Criteria for higher volumes of CCP were at the assistant physician’s discretion but included mechanical ventilation and/or absence of clinical improvement within 2 days from the first CCP transfusion. CCP units with nAbs titers &[gt]160 were transfused, except for the first eleven patients included in the first two weeks of the study, when nAbs from CCP units were not available before transfusion, comprising sixteen CCP doses, with nAb geometric mean titer ranging from 20 to 640. No other interventions on supportive COVID-19 treatment were made by the research staff. Clinical and laboratory information were collected until discharge: baseline demographic data, comorbidities, laboratory parameters, days of illness onset before hospital and ICU admission, other COVID-19 supportive therapies and mechanical ventilation. Titration of nAbs was performed using the cytopathic effect-based virus neutralization test (CPE-based VNT) with SARS-CoV-2 (GenBank MT126808.1) [16] with patient samples on the day of enrolment (day 0) (nAbs P0) and on day 5 (nAbs P5) after CCP transfusions. Clinical status of patients was assessed according to severity organ failure assessment (SOFA) score on day 0 and evolution was assessed with adapted World Health Organization (WHO) ordinal scale [17] (table 1) on days 0, 5, and 14. Intensive care unit (ICU) and hospital length of stay, duration of mechanical ventilation, and disease evolution were registered.

**Table 1.** Demographic and Clinical data of studied patients

|  |  |
| --- | --- |
| Gender |  |
| Female | 30 (28.8%) |
| Male | 74 (71.2%) |
| Total | 104 (100.0%) |
| Age (years) |  |
| Mean (SD) | 64 (15) |
| Min- Max | 31-95 |
| ABO group |  |
| A | 45 (45.2%) |
| O | 44 (42.3%) |
| B | 10 (9.6%) |
| AB | 3 (2.9%) |
| Comorbidities (%) | 93 (89.4%) |
| Hypertension | 54 (51.9%) |
| Diabetes mellitus | 31 (29.8%) |
| Obesity | 36 (34.6%) |
| Use of other supportive therapies n (%) | 75 (72.1%) |
| Hydroxychloroquine | 34 (33.7%) |
| Azythromicin | 72 (70.2%) |
| Immunoglobulin | 3 (2.9%) |
| Tocilizumab | 7 (6.7%) |
| Mechanical ventilation | 51 (49%) |
| Transfusion of CCP |  |
| 300 ml | 19 (18.8%) |
| 400 ml | 23 (22.8%) |
| 600 ml | 59 (58.4%) |
| In hospital length of stay –days (n=61) |  |
| Median (1<sup>o</sup> IQR, 3<sup>o</sup> IQR) | 23 (14-33) |
| Min- Max | 4-117 |
| ICU length of stay-days (n=57) |  |
| Median (1<sup>o</sup> IQR, 3<sup>o</sup> IQR) | 13 (7-21) |
| Min- Max | 2-65 |
| Duration of mechanical ventilation (n=36) |  |
| Median ((1<sup>o</sup> IQR, 3<sup>o</sup> IQR) | 10 (5-16) |
| Min-Max | 3-61 |
| nAbs P0 (n=98) |  |
| Median (1<sup>o</sup> IQR, 3<sup>o</sup> IQR) | 160 (20-640) |
| Min- Max | 10-1240 |
| nAbs P5 (n=93) |  |
| Median (1<sup>o</sup> IQR, 3<sup>o</sup> IQR) | 5120 (640-5120) |
| Min- Max | 10-5120 |
| nAbs T (nAbs mean geometric titer) |  |
| Median ((1<sup>o</sup> IQR, 3<sup>o</sup> IQR) | 403.2 (254-640) |
| Min-Max | 20-3044.4 |

\*\*: 100 unit increase; #500 unit increase; nAbsP0: Neutralizing antibody titers from patient samples before convalesce plasma transfusion; nAbsP5: Neutralizing antibody titers from patient samples collected at day 5 after CCP transfusion, nAbsT: geometric mean titer of CCP units transfused

**Table 2.**
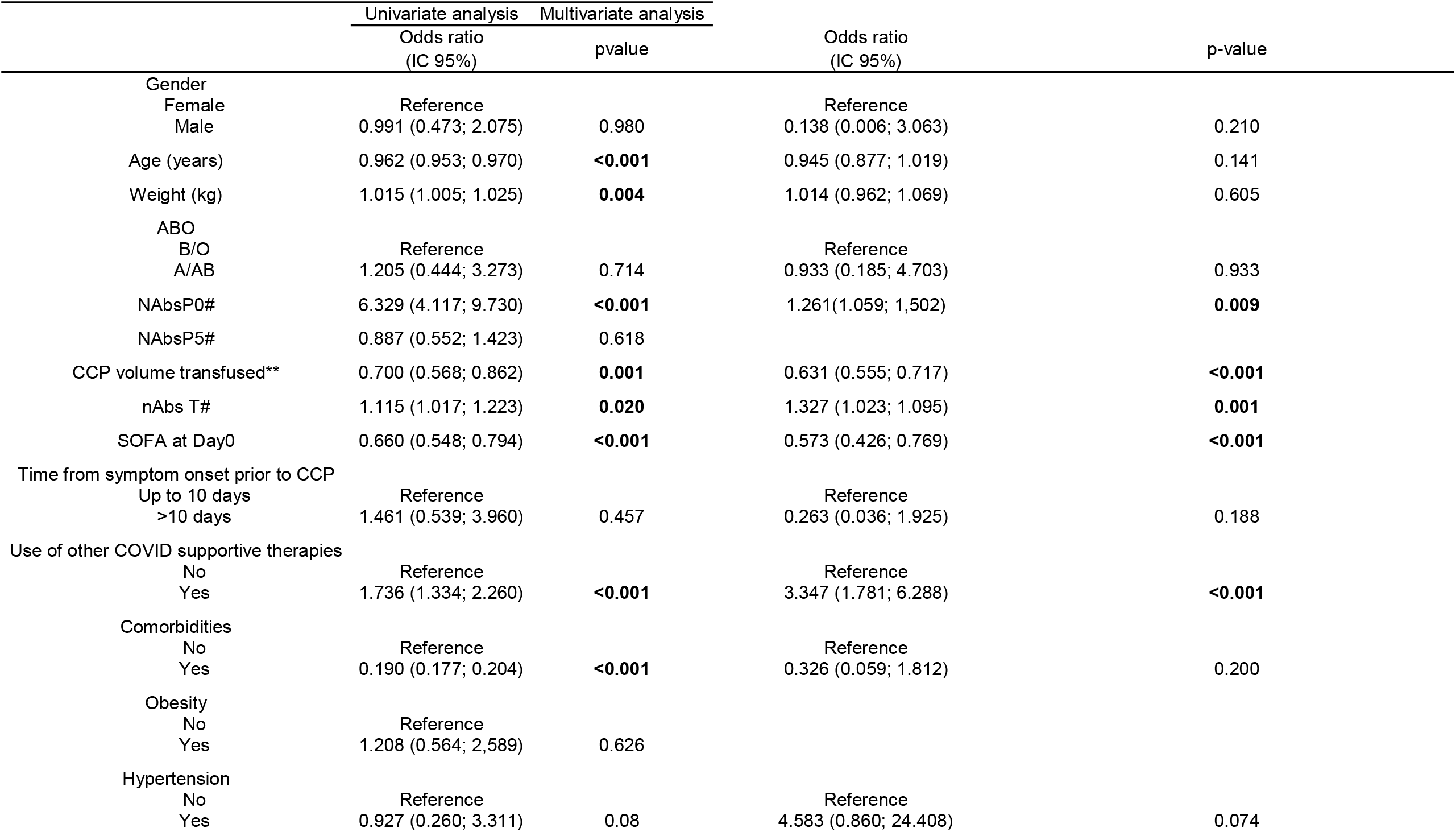
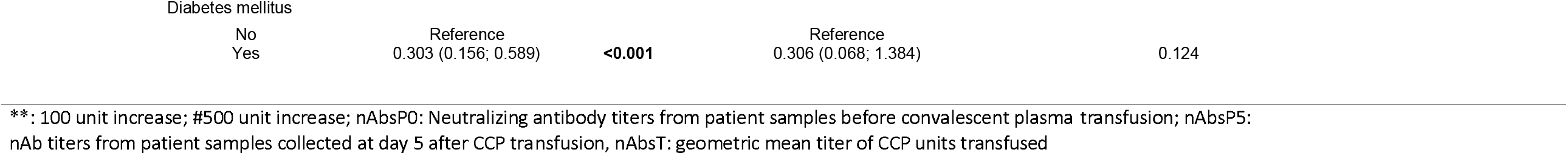
Clinical improvement on Day 14 post-CCP transfusion

**Table 3.** Duration of mechanical ventilation post-CCP transfusion

|  | Univariate analysis |  | Multivaried analysis |  |
| --- | --- | --- | --- | --- |
|  | Mean ratio<br>(CI 95%) | - p value | Mean ratio<br>(IC 95%) | - p value |
| Gender |  |  |  |  |
| Female | Reference |  | Reference |  |
| Male | 1.239 (0.681; 2.257) | 0.483 | 1.226 (0.631; 2.379) | 0.548 |
| Age (years) | 0.996 (0.988; 1.004) | 0.288 | 0.978 (0.963; 0.992) | **0.003** |
| Weigth (kg) | 0.996 (0.988; 1.004) | 0.332 | 0.992 (0.989; 0.994) | **<0.001** |
| ABO |  |  |  |  |
| B/O | Reference |  | Reference |  |
| A/AB | 0.908 (0.453; 1.818) | 0.785 | 0.967 (0.343; 2.730) | 0.950 |
| nAbs P0# | 0.648 (0.523; 0.803) | **<0.001** | 0.961 (0.951-0.972) | **<0.001** |
| nAbs P5# | 1.005 (0.942; 1.073) | 0.869 |  |  |
| Volume transfused** | 1.204 (1.010; 1.436) | **0,038** | 0.933 (0.697; 1.248) | 0.639 |
| Mean geometric titer of transfused nAbs (NAbsT)# | 0.941 (0.865; 1.025) | 0.164 | 0.784 (0.434; 1.417) | 0.420 |
| SOFA score at D0 | 1.165 (1.151; 1.180) | **<0.001** | 1.180 (1.142; 1.221) | **<0.001** |
| Time from disease onset until CCP transfusion |  |  |  |  |
| Up to 10 days | Reference |  | Reference |  |
| >10 days | 1.170 (0.514; 2.659) | 0.709 | 1.105 (0.829; 1.473) | 0.494 |
| Use of other supportive therapies |  |  |  |  |
| No | Reference |  | Reference |  |
| Yes | 0.378 (0.261; 0.548) | **<0.001** | 0.544 (0.287; 1.031) | 0.062 |
| Obesity |  |  |  |  |
| No | Reference |  |  |  |
| Yes | 1.155 (0.678; 1.967) | 0.596 |  |  |
| Hipertension |  |  |  |  |
| No | Reference |  | Reference |  |
| Yes | 0.963 (0.536; 1.729) | 0.899 | 0.977 (0.574; 1.664) | 0.932 |
| Diabetes mellitus |  |  |  |  |
| No | Reference |  | Referencia |  |
| Yes | .,342 (1.065; 1.691) | **0.013** | 0.906 (0.557; 1.472) | 0.690 |

\*\*: 100 unit increase; #500 unit increase; nAbsP0: Neutralizing antibody titers from patient samples before convalesce plasma transfusion; NAbsP5: Neutralizing antibody titers from patient samples collected at day 5 after CCP transfusion, NAbsT: geometric mean titer of CCP units transfused

**Table 4.**
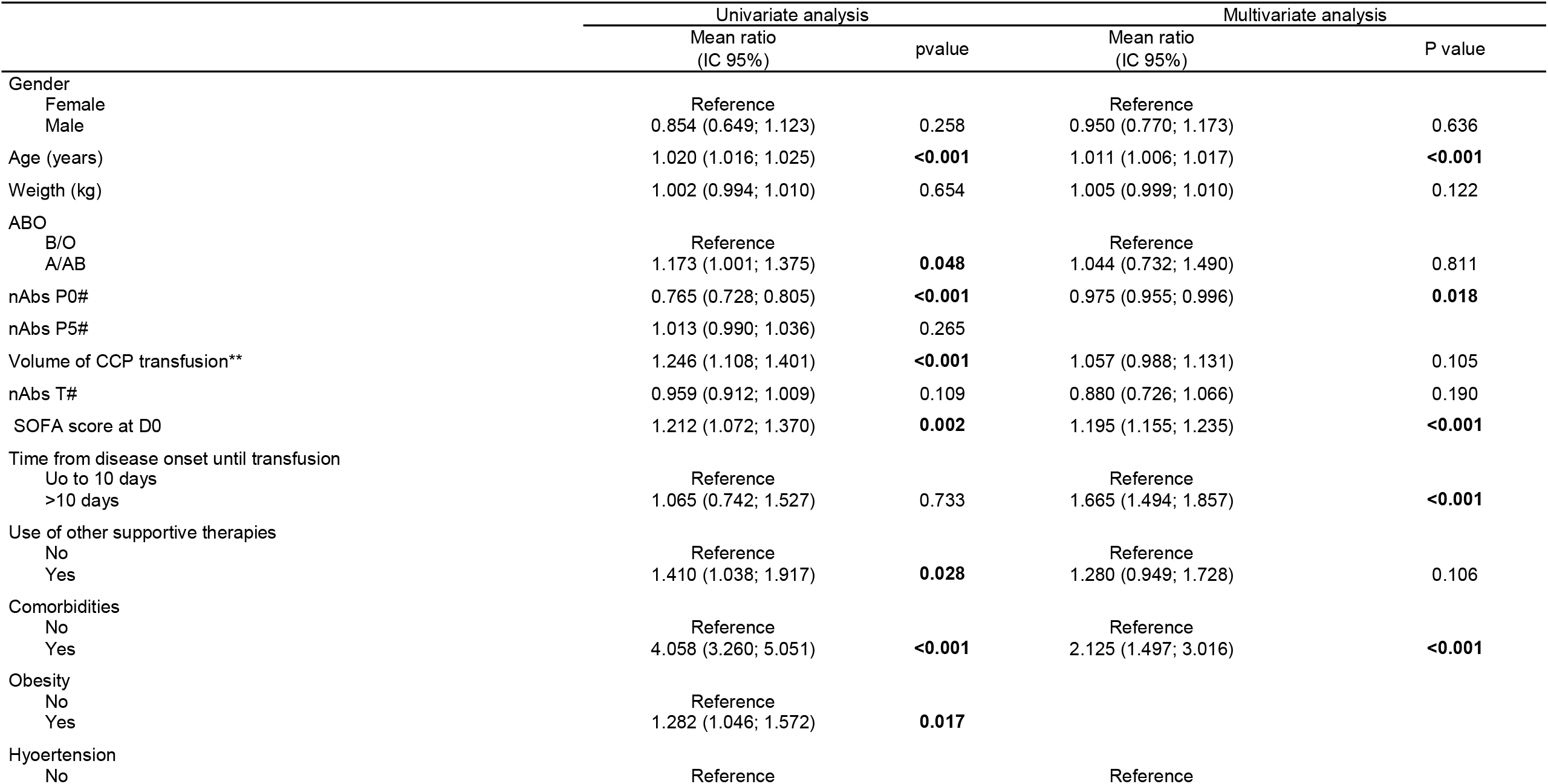
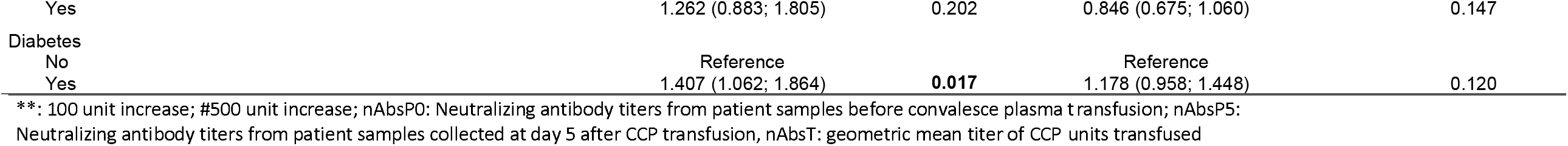
ICU length of stay post-CCP transfusion

### Outcomes

The primary outcome was a clinical improvement on day 14, defined as a reduction of at least two points on an adapted WHO ordinal scale. Secondary outcomes were intensive care unit length of stay (ICU LOS) and duration of invasive mechanical ventilation.

### Statistical Analysis

Baseline characteristics were described as counts and percentages for categorical variables or mean and standard deviation (SD) for continuous variables with a normal distribution. Median and interquartile range (IQR) were used to describe continuous variables with asymmetrical distribution. Univariate and multivariate regression models were adjusted to assess the outcomes: intensive care unit (ICU) length of stay, duration of mechanical ventilation, and clinical improvement on day 14 (binary). Explanatory variables considered were: mean geometric titer of Nabs from CP units transfused (NAbsT), neutralizing antibody titers from patients before transfusion (NAbsP0), neutralizing antibody titers from patients on Day 5 (NAbsP5), the total volume of CP transfused, age, gender, weight, ABO group, severity organ failure assessment score on admission day (SOFA D0), use of other supportive therapies for COVID-19 (azithromycin, hydroxychloroquine, steroids, tocilizumab, human immunoglobulin, antiviral therapies), and comorbidities. To analyze if earlier CP transfusion had an impact on outcomes, we also analyzed if CP transfusion was performed up to 10 days of disease onset or later. For each outcome, we performed a generalized estimating equations model to account for the dependence of data collected at each of the three institutions. For mechanical ventilation, we used negative binomial distribution and for ICU LOS we used gamma distribution, both with log link functions. Clinical improvement on day 14 was assessed with a binomial distribution and logit link function. Results were presented as mean ratios (MR) or odds ratios (OR) with 95% confidence intervals and p values, p&[lt]0.05 was considered statistically significant. For the analysis, we used IBM SPSS Statistics for Windows, Version 24.0. 2016, R package [18], and ggplot2 [19].

## Results

Mean age of patients was 64 years (SD: +/- 15 yrs), 74 patients (71.1%) were male, and 93 (89.4%) had different comorbidities - arterial hypertension was present in 51.9% of the individuals, diabetes mellitus in 29.8% and 34.6 % were obese. The median body weight was 84 kg. Regarding the ABO blood group, 45 (45.2%) were A individuals, 44 (42.3%) were O, 10 were B (9.6%) and 3 (2.9%) were AB. The vast majority of patients (82.7%) also received other therapies (azithromycin, hydroxychloroquine, tocilizumab and/or combinations). Fifty-one patients (49%) required invasive mechanical ventilation. NAbs from patients on day 0 (immediately before CCP transfusions - NAbsP0) varied from 10 to 1240 [1^st^ IQR 640-3 ^rd^ IQR 5120]. To consider the dose-effect of nAbs on each outcome, the total amount of nAbs were calculated using geometric mean nAbs titer from CCP units (NAbsT), which varied from 20 to 3044 [1^st^ IQR 254-3^rd^ IQR 640]. Three patients received doses of 200 ml (2.9%), nineteen patients received doses of 300 ml (18.2%), 23 patients received 400 ml (22.8%) and 59 received 600 ml (56.7%) (Table 1). CCP transfusions were well-tolerated and 5 (4.8 %) mild to moderate transfusion reactions occurred: 3 febrile non-hemolytic reactions, 2 allergic reactions. Two suspected cases were screened for TRALI, with negative results. Of note, only plasma from male, nulliparous women, or women with a history of up to 2 gestations and negative HLA screening (Lifecodes lifescreen Deluxe – Immucor) were suitable for CCP donation. No severe adverse events were observed. Six (5.7%) had thromboembolic events (TE) after CCP infusion, even under thrombotic prophylaxis or on full anticoagulation due to TE diagnosed before the enrolment. These rates are somewhat lower than the rates observed in hospital COVID patients in general [20,21]

Figure 1 shows the timeline of clinical evolution after the onset of illness (OIL) for the 104 patients. Median times from OIL to hospital admission was 6.5 days [1^st^ IQR 3-3 ^rd^ IQR 9.9], and from OIL to dyspnea was 6 days [IQR 3-8]. Median times from OIL to ICU admission was 8.7 days [IQR 5.4-11.0] and from OIL to CCP transfusion was 10 days [1 ^st^ IQR 8.0-3rd IQR 13.0].

**Figure 1.**
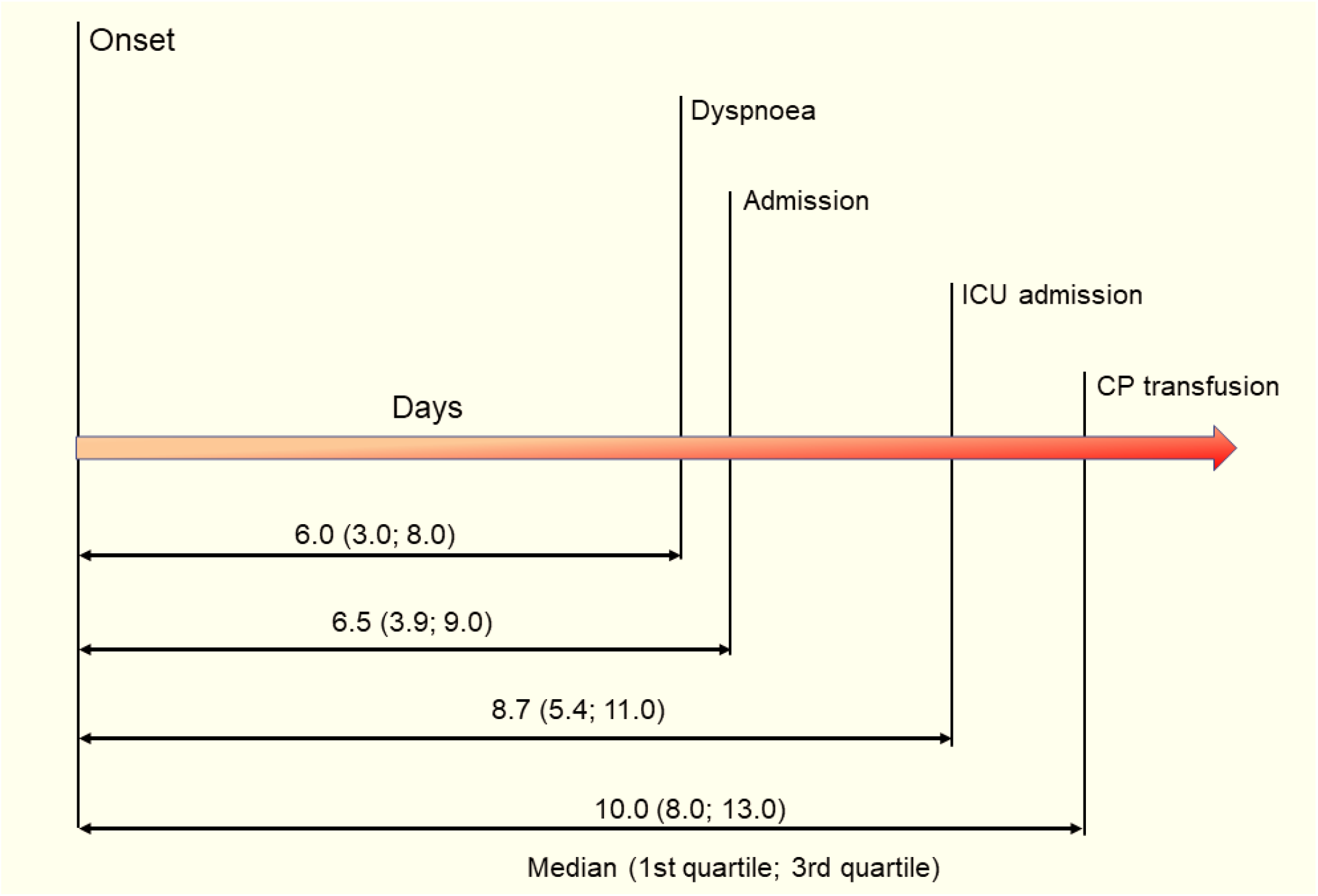
Timeline of COVID-19 patients included in the study

Clinical improvement was assessed by the World Health Organization scale score at days 0, 5 and, 14. By day 5, 19 (18.2%) patients have achieved clinical improvement. On day 14, the proportion of patients who had improved raised to 41%, with 49.4% of patients with mild disease (WHO ordinal scale between 0-4) and 38 (36.5%) had already been discharged. There were eleven deaths (10.5%), and one event happened before day 7 after CCP transfusion (figure 2).

**Figure 2.**
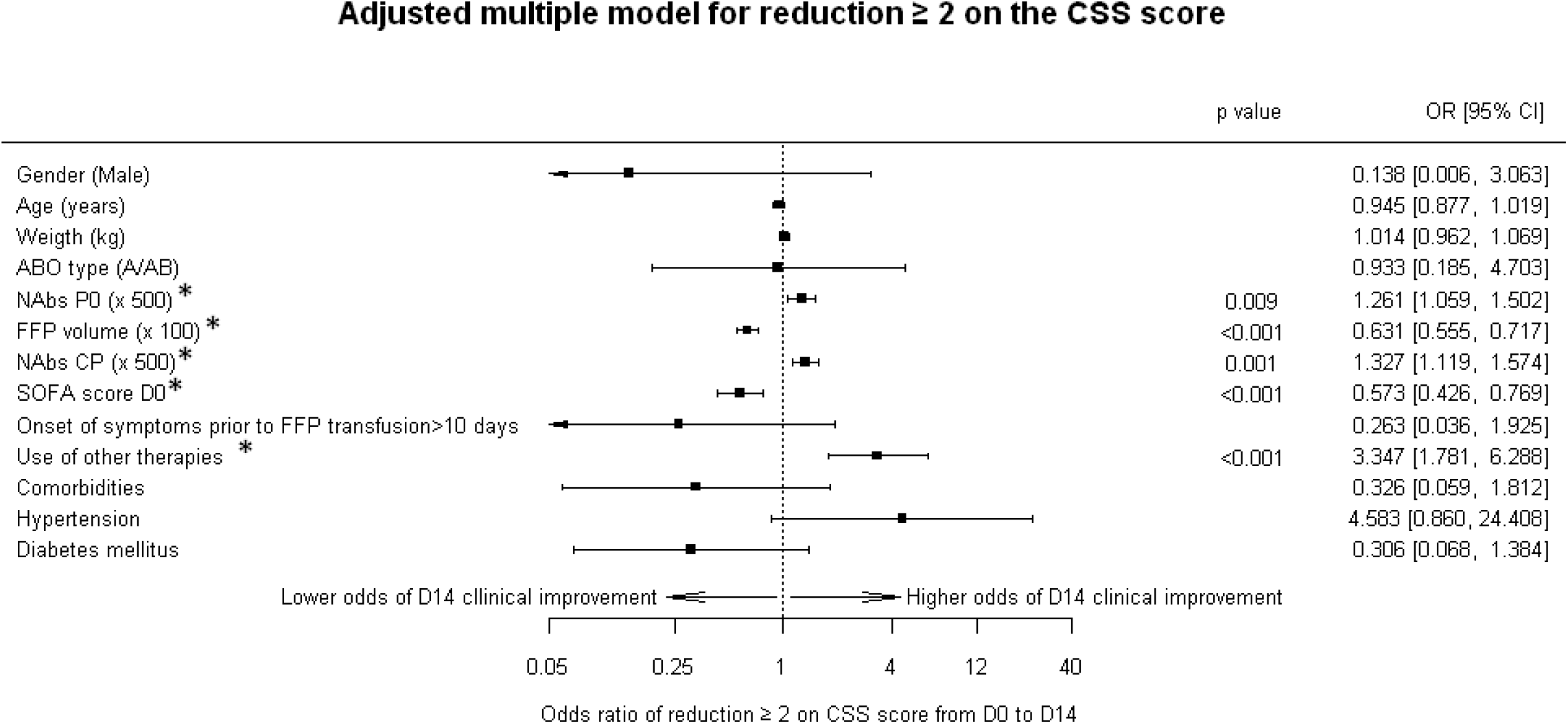
Clinical Improvement on Day 14 post-CCP transfusion. Obs. Left side means worse performance, right side means better performance. *Means had statistical significance Nabs P0 = samples patients neutralizing antibodies measured on day of enrolment; FFP volume = fresh frozen plasma volume; nAbs CP = neutraziling antibodies from the convalescent plasma. Variables without a p value means p was non-significant.

**Figure 3.**
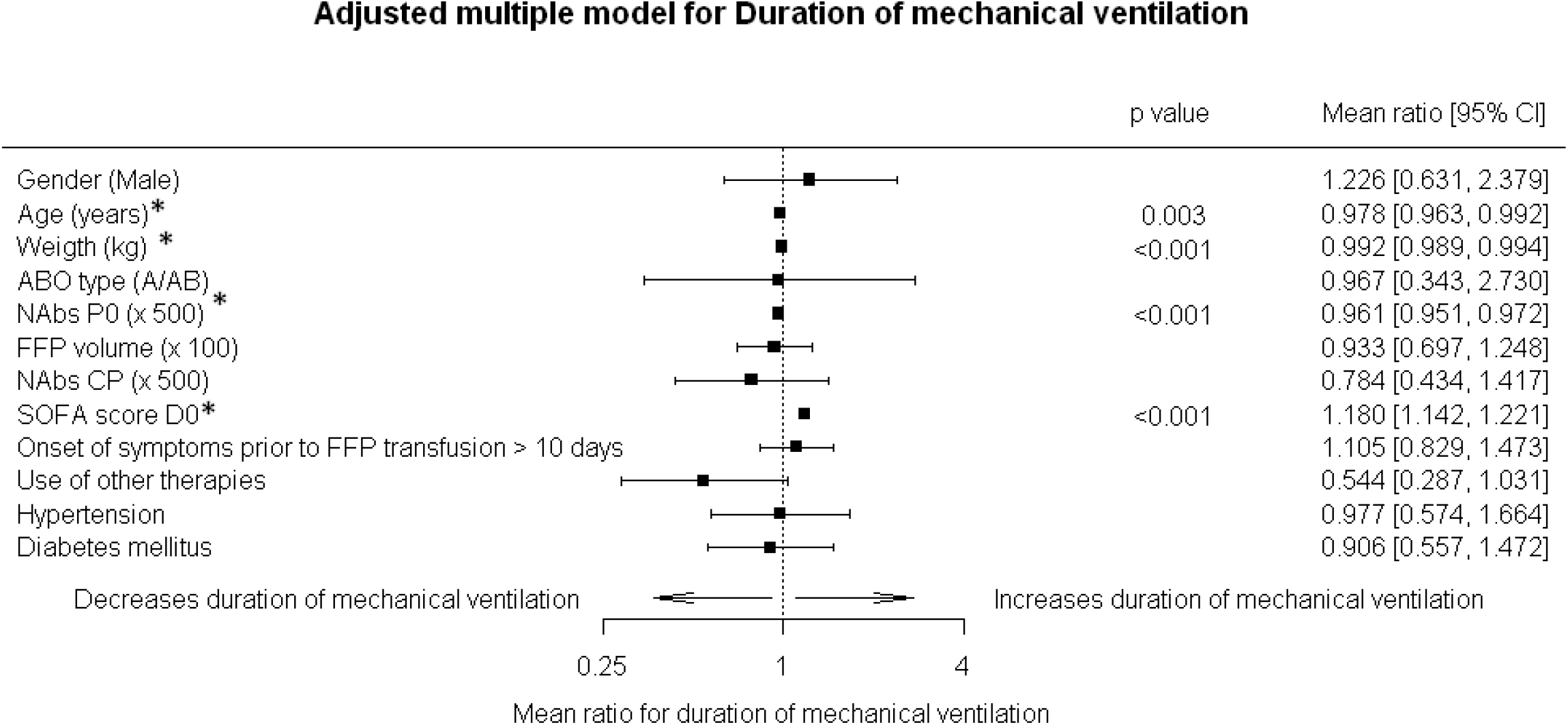
Duration of Mechanical Ventilation post-CCP transfusion. Obs. Left side means worse performance, right side means better performance. *Means had statistical significance. Nabs P0 = samples patients neutralizing antibodies measured on day of enrolment; FFP volume = fresh frozen plasma volume; nAbs CP = neutraziling antibodies from the convalescent plasma. Variables without a p value means p was non-significant.

**Figure 4.**
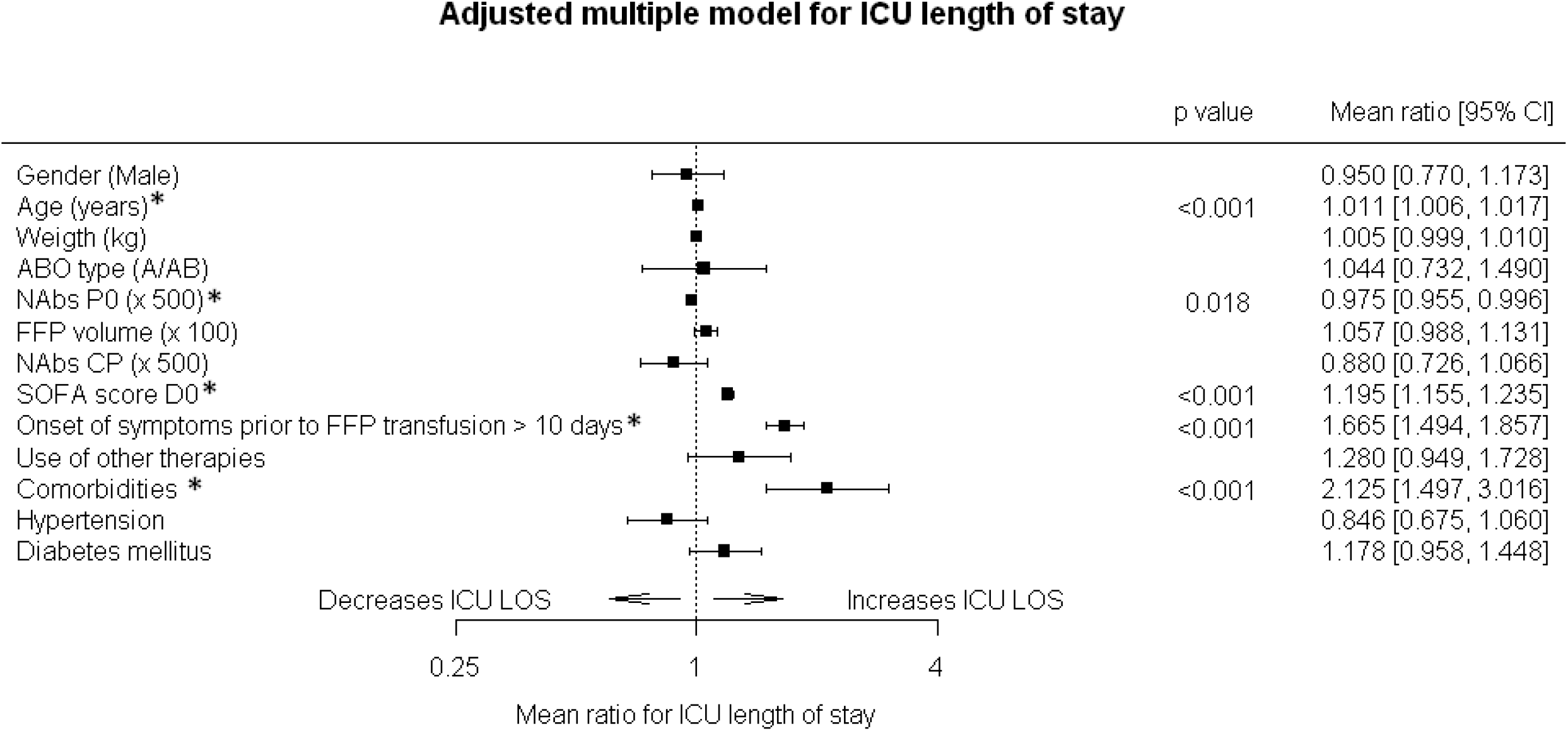
ICU Length of Stay post-CCP transfusion. Obs. Left side means worse performance, right side means better performance. *Means had statistical significance nAbs P0 = samples patients neutralizing antibodies measured on day of enrolment; FFP volume = fresh frozen plasma volume; nAbs CP = neutraziling antibodies from the convalescent plasma. Variables without a p value means p was non-significant.

**Figure 5.**
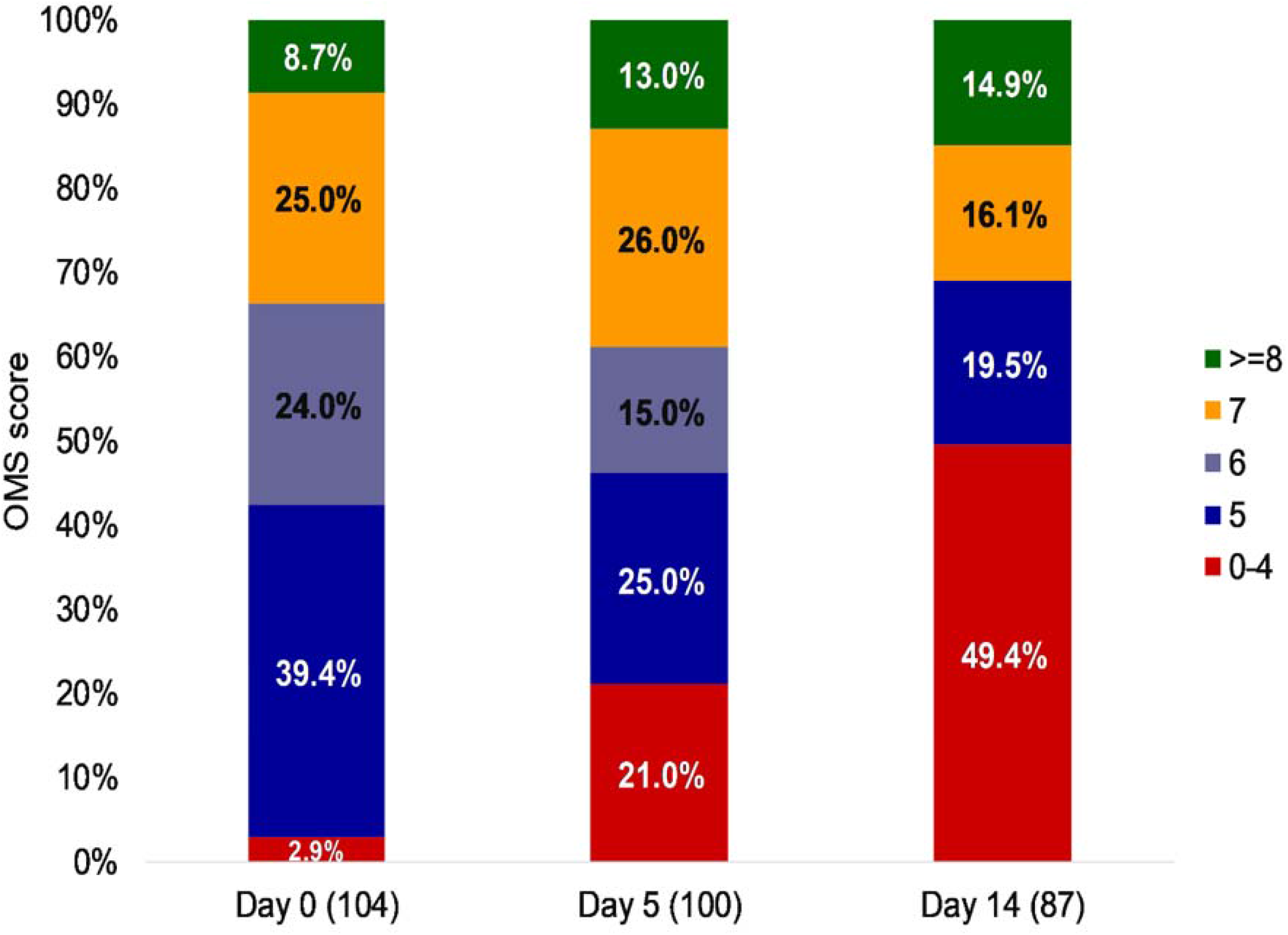
Adapted from OMS Score on days 0, 5, and 14 post-CCP transfusion

Multivariable analysis showed that circulating nAbs developed by the patients before transfusion (NAbsP0) (OR=1.261; CI95%: 1.059-1.502, p=0.009), mean geometric nAbs titer from transfused CCP units (NAbsT) (OR=1.327; CI95%: 1.119-1.574, p=0.001) and use of other supportive therapies for COVID-19 (OR=3.347; CI95%: 1.781-6.288, p< 0.001) are associated with higher odds of clinical improvement on day 14. On the contrary, SOFA score on day 0 (OR=0.573; CI95%: 0.426-0.769, p<0.001) and total CCP volume transfused (OR= 0.631; CI95%: 0.555-0.717, p< 0.001) were associated with lower chances of clinical improvement. Age, gender, ABO group, time from onset of illness until CCP transfusion, comorbidities, and exposure to other therapies had no impact on 14-day clinical improvement.

We sought to determine predictive variables for the duration of mechanical ventilation and ICU LOS. Duration of mechanical ventilation was available for 36 patients. Data from 11 patients who died and 4 patients who have not been discharged at the end of the analysis were not considered for this evaluation. Mechanical ventilation support varied from 3 to 61 days, median duration was 10 days [1^st^ IQR 5-3^rd^ IQR 16]. Logistic regression models were performed to identify predictive variables. Multivariable analysis showed that level of patient nAb titers prior to transfusion (NAbsP0) (*Mean ratio* MR=0.961; CI95%:0.951-0.972, p<0.001), SOFA score on day 0 (MR=1.180; CI95%: 1.142-1.221, p<0.001), age (MR= 0.978; CI95%: 0.963-0.992, p<0.003) and body weight (MR=0.992; CI95%:0.989-0.994, p<0.001) were independently associated to duration of mechanical ventilation support. The higher the NAbsP0 before CCP infusion, the shorter the need for mechanical ventilation support. Paradoxically to what has been described in the medical literature [22,23,24], older patients and more obese were also associated with shorter durations of invasive mechanical ventilation. The severity of patients assessed with SOFA score also revealed associations with longer periods of mechanical ventilation support. Other variables failed to provide associations with the length of mechanical ventilation support.

We also analyzed predictive variables for ICU LOS. Multivariate logistic regression showed that higher NAbsP0 (MR=0.975; CI95%: 0.955-0.996, p<0.018) were associated with shorter periods of ICU LOS. Conversely, time for transfusion after 10 days of symptom onset (MR=1.665; CI95%: 1.494-1.857, p<0.001), presence of comorbidities (MR=2.125; CI95%: 1.497-3.016, p< 0.001), severity of the disease on day 0 (MR=1.195; CI95%:1.1155-1.235, p<0.001) and age (MR=1.011; CI 95%: 1.006-1.017) were associated with longer periods in ICU. NAbs T showed no statistical significance on reduction of ICU LOS (MR= 0.880; CI95%: 0.726-1.066, p=0.190)

## DISCUSSION

We report our experience with CCP transfusion in 104 severe critically ill patients with COVID-19. The first data drawn from this study is that CCP is a safe procedure, where no major hazard effects were observed in this group of severe patients. CCP transfusion is based on the fact that it provides nAbs able to reduce the viral burden and prevent systemic manifestations in susceptible patients [25]. Several observational studies reported data favoring the efficacy of convalescent plasma [26, 27, 28]. Conversely, recently published randomized clinical trials on convalescent plasma for COVID-19 [29,30,31] showed no benefit on mortality or clinical improvement on day 30 [32]. Also, a systematic review analyzing CCP and hyperimmune immunoglobulin failed to provide robust evidence regarding the clinical efficacy of both products [33], highlighting the need for more research on this field.

We analyzed predictive factors for clinical improvement on day 14. Forty-one patients achieved clinical improvement, and multivariable logistic regression showed that nAbs T (p= 0.001), nAbs P0 (p=0.009), and use of other supportive therapies (p<0.001) were statistically associated with higher odds of clinical improvement. At each −500 unit increase in nAbs T, the odds of clinical improvement raised by 32.7%. Median geometric nAbs was 403.2, interquartile range 254-640. Our findings differ from previous studies on CCP efficacy. A randomized controlled trial published by Gharbharan *et al*,, although prematurely halted, showed no difference (p = 0.58) on improvement in disease severity by day 15. The PLACID trial [30] also failed to provide evidence in favor of CCP transfusion, once plasma was not associated with a reduction in progression to disease severity at 28 days. No significant differences in clinical improvement or overall mortality were observed by Simonovitch *et al*,[32], who transfused CCP units with median nAbs titer similar to ours (median nAbs: 300, interquartile range 136-511). It is still unknown if higher titer nAbs would have resulted in better outcomes. We acknowledge that CCP transfusion impacts on clinical improvement as a modest effect (OR 1.327, CI95% 1.119-1.574 p: 0.001), as opposed to the impact of using other COVID 19 supportive therapies (OR 3.347 CI95% 1.781-6.288, p<0.001). More research is still needed to better clarify the benefit of CCP in clinical improvement.

Considering ICU LOS and length of mechanical ventilation, in our analysis, nAbs P0, but not nAbsT, were associated with a significant reduction in ICU LOS (p=0.018) and duration of mechanical ventilation (p<0.001). At each −500 unit increase on the mean geometric titer of nAbsP0, a 2.5% reduction on ICU LOS was expected and a 3.9% reduction on the duration of mechanical ventilation. On the contrary, nAbs T had no impact on these same outcomes. Our findings reinforce the fact that patient nAbs and not nAbs from transfused CCP units may determine the outcome in severe disease. Accordingly, Wang [34] showed that patients with more severe symptoms tended to have higher antibody titers. Hence, CCP transfusion might not add benefit in this scenario, and antibody screening before CCP transfusion could be useful to identify potential individuals who could benefit from this passive therapy [29]. This is important especially on critically ill patients, reported having solid antibody response. In our study, data about the nAbs status of patients were not available before CCP transfusion and such analysis was not possible.

Also, our findings show that timely administration of CCP is relevant for clinical outcomes. Administration of CCP after 10 days of symptom onset was associated with increases in ICU length of stay in a statistically significant manner (p<0.001). Antibody responses to SARS-CoV-2 seem to appear between 2 −3 weeks after initiation of symptoms [35] and nAbs specifically reach their peak within 10-15 days after disease onset [36]. In a prematurely interrupted randomized trial, transfusion of CCP in later stages did not result in significant clinical improvement within 28 days compared to standard treatment [31]. In this study, the median time from disease onset to CCP intervention was 30 days, twice the period from ours. On the other hand, in a case-control study published by Liu *et al*, [2], CCP transfusion was administered with a median time of 7 days from admission to transfusion. Improvements in survival in patients not on mechanical ventilation support and decreased oxygen requirements were observed. Taken this study with ours are following the fact that earlier initiation of passive immunotherapy might provide better outcomes. [2,27, 37,38,39].

We also observed that the severity of the disease at day 0 (SOFA D0), age, and comorbidities, in general, was associated with increases in ICU length of stay. However, these findings were not reproducible in the duration of mechanical ventilation. Our data partially differs from Gamberini *et al*,, who demonstrated that age, SOFA score at ICU admission, PaO_2_/FiO_2_, renal and cardiovascular complications, and late-onset VAP were all independent risk factors for prolonged mechanical ventilation in patients with COVID-19 [40]. Differences found may be due to the low number of patients analyzed and different methodologies implemented.

Despite the study limitations such as the size of the cohort and the fact that it is a one-arm prospective study, our data have shown an association between patients’ previously acquired nAbs and clinical outcomes. Our analysis also suggests that nAbsT was associated with clinical improvements on day 14. Besides, nAbsP0 but not nAbsT have an impact on ICU LOS and duration of mechanical ventilation. The potential value of timely administration of CCP transfusion before day 10 of disease onset on the improvement of clinical outcomes was also demonstrated. In conclusion, we consider these data are useful parameters to guide future CPP transfusion strategies to COVID-19.

## Data Availability

All related data will be available under special requitrement to the corresponding author

## Acknowledgements

The authors would like to thank EV Souza for the statistical analysis and advise, also to the following physicians for referring their patients to this study: ALP Albuquerque, AC Nicodemo, D Deheinzelin, E Negri, LFL Carvalho, LF Cardoso, MAR Cuoco, P Seferian-Junior, R Kairalla, T Pfiffer, Y Novis, H Souza, T Zinsly, A Pacheco, G Johanson, FT Moreira, S Santoro, B Normandia, RM Carraro, E Meyer, A Lichtentstein, F Bacal, M Amar, CAF Santos, D Gondenberg, A Jaime, M Rodrigues-Junior, S Filizola, H Bacha, H Bogossian, C Hoelz, M Erlichman, E Pfefferman, R Kondo, MH Kuwakino, R Peres, GL Bub, DS Levi and FG Menezes.

## Author contribution

Conceptualization: JMK, SW, APHY, RMF, CBB; investigation: JMK, SW, APHY, RMF, CBB, GC, PS, RA, MAB, RRGM, DBA, ELD.; formal analysis: JMK, SW, APHY, RMF, CBB, ARM; resources: LFLR, LVR writing: APHY, SW, JMK, CBB.

